# Prevalence of visual impairment and its causes in adults aged 50 years and older: Estimates from the National Eye Surveys in Malaysia

**DOI:** 10.1101/2024.02.18.24303005

**Authors:** Mohamad Aziz Salowi, Nyi Nyi Naing, Norasyikin Mustafa, Wan Radziah Wan Nawang, Siti Nurhuda Sharudin, Nor Fariza Ngah

## Abstract

**Background:** Population surveys are necessary to measure a community’s eye care needs. We conducted simultaneous surveys in two regions in Malaysia in 2023 to estimate the prevalence of visual impairment (VI), identify its main causes, and compare the results with the survey in 2014.

**Methods:** The surveys were simultaneously done in Eastern and Sarawak administrative regions using the Rapid Assessment of Avoidable Blindness (RAAB) technique. It involved a multistage cluster sampling method, each cluster comprising 50 residents aged 50 years and older. The prevalence of VI (blindness, severeness, moderate, and early) and its primary cause were determined through a visual acuity check and eye examination with a hand-held ophthalmoscope. Results were compared with the previous survey in 2014.

**Results:** A total of 10,184 subjects were enumerated, and 9,709 were examined (94.5% and 96.2% responses for Eastern and Sarawak, respectively). The prevalence of blindness and severe VI appeared to decrease. For blindness: Eastern 1.4%, 95%CI (0.9, 1.9) to 0.8%, 95%CI (0.5, 1.1) and Sarawak: 1.6% 95%CI (1.0, 2.1) to 0.6%, 95%CI (0.3, 0.9). For severe VI: Eastern 1.2%, 95%CI (0.8, 1.7) to 0.9%, 95%CI (0.6, 1.1) and Sarawak 1.1% 95%CI (0.6, 1.6) to 0.9%, 95% CI(0.6, 1.2). The main cause of blindness was untreated cataracts: 77.3% (Eastern) and 75.0% (Sarawak). Diabetic retinopathy was the 2^nd^ main cause of blindness for Eastern at 9.1%, but it only caused early to severe VI in Sarawak.

**Conclusion:** The prevalence of blindness and severe VI were reduced. The reduction could have been attributed to a community cataract program implemented soon after the survey in 2014. However, more efforts are needed to address the high percentage of avoidable blindness within both regions.

## Introduction

Malaysia is one of the 37 member states of the World Health Organisation (WHO) Western Pacific Region [1]. As part of the global and regional eye health agendas, Malaysia has been actively engaged with other countries within the Western Pacific Region in planning, implementing, and monitoring community programs related to the Prevention of Blindness and Low Vision (PBL) initiatives. The activities’ coverage expanded and strengthened after the country endorsed and signed the World Health Assembly (WHA) 66.4 resolution in May 2013 [2,3]. By signing the resolution, Malaysia is committed to being part of the Global Action Plan, the core activity and aim of which is providing Universal Health Care (UHC) to the population [3].

Malaysia is divided into six regions for the PBL administrative purposes: Northern, Eastern, Central, Southern, Sabah and Sarawak (Fig 1). Simultaneous surveys were done in 2014 for each region as a baseline, and the results were published. The main finding of the surveys was a higher prevalence of blindness in the peripheral regions, namely Sabah, Sarawak, Northern, and Eastern regions, and a discrepancy between all regions [4]. One of the national action plans planned and implemented was to introduce a community interventional eye care program, which could potentially reach the underserved population especially in remote areas. Guided by the pre-existing basic community program and the availability of eye-trained human resources, the Eastern Region and Sarawak were selected as the regions where this program would be piloted.

**Fig 1.**
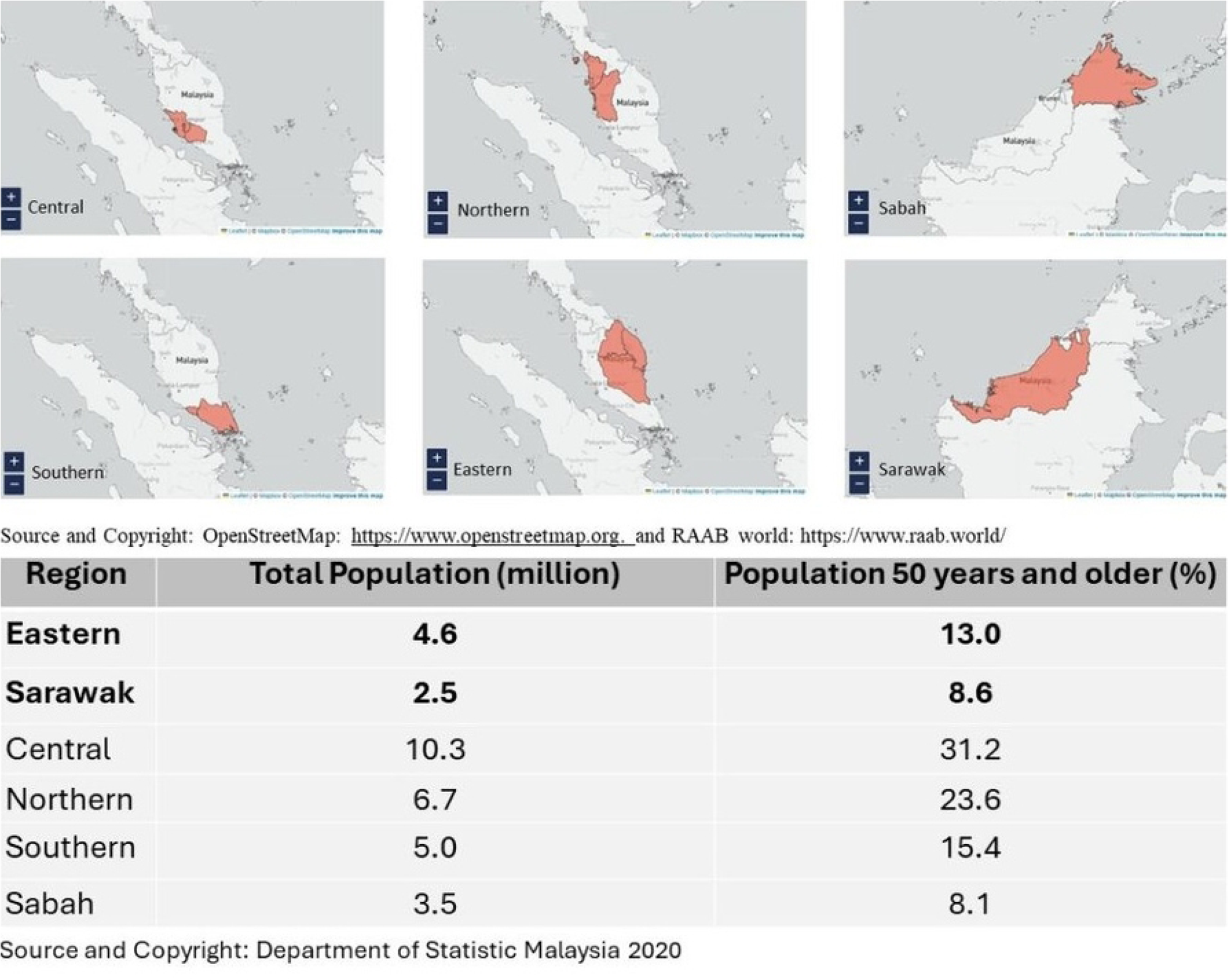
Survey administrative regions, total population, and percentage of individuals 50 years and older in Malaysia. (This study surveyed Eastern and Sarawak only).

This study aims to determine the difference or improvement in the prevalence of visual impairment (VI) between the two administrative regions from the previous survey.

## Material and Methods

A follow-up survey was required to compare the prevalence of VI before and after the community eye care program’s implementation. These cross-sectional, population-based surveys, which followed the World Health Organization (WHO) RAAB protocol, were conducted simultaneously from 27 July 2023 to 7 October 2023. The surveys also collected data on cataract surgical coverage (CSC) and cataract surgical outcomes, the results of which will be discussed in another manuscript.

Each region had six (6) data collector teams comprising three persons: two doctors and one allied health staff member trained in ophthalmology. Each team was responsible for surveying 16-17 randomly selected clusters, examining 50 residents aged 50 years and older. Population sampling was per RAAB methodology, a widely used, WHO-recommended method for population-based surveys of the prevalence of VI and its causes. The RAAB survey protocol and methodology have been described elsewhere [5].

### Sampling frame

Department of Statistics, Malaysia (DOSM) conducts nationwide data collection for the National Population and Housing Census once every ten years. An Enumeration Block (EB), the smallest population unit, with 80-120 living quarters each, is demarcated based on the latest findings and the population distribution, followed by the development of a corresponding geographical map, indicating the exact location and boundaries of each EB. The EBs are gazetted for field work operations, for example, Morbidity, Nutrition, Household Expenditure, and Labor Force Survey [6]. The complete list of all EBs from the 2020 national census was used to select clusters for the RAAB. A total of 105 EBs were randomly chosen for Eastern and 98 EBs for Sarawak regardless of strata, using the Probability Proportionate to Size (PPS) technique. Individual EB codes and the corresponding maps were then used to identify the location of the EBs during fieldwork data collection.

### Training

Training for survey teams was conducted separately in each region before the fieldwork by a certified Western Pacific RAAB trainer to ensure data quality and strict adherence to study protocol. Survey team members were required to attend four training days, including RAAB lectures, inter-observer variation assessment and an actual survey in one of the nearby EBs during fieldwork. Each region had one coordinator responsible for the smooth implementation and progress of the survey.

### Survey Methods

Each team was assigned to survey 16-17 EBs. Subjects were selected from each block using the Compact Segment Sampling method. The population area was divided into segments of equal population size, enough to provide the required number of eligible people aged 50 years and older. The survey started in a randomly selected segment until 50 eligible people had been examined. If all houses in the segment had been visited and the number of subjects was insufficient, then a second random segment was picked for the survey team to continue until they recruited 50 subjects.

If the subject was unavailable at home, the contact number would be taken from the neighbours, and a revisit would be done before the team left the survey area. If the subject could not be examined after three revisits, this person would be recorded as ‘Not Available’, and the vision status reported by relatives or neighbours would be taken. Door-to-door interviews were conducted in each randomly selected EB. Subjects were recruited if they were 50 years and older, a Malaysian resident of at least six months, and gave informed consent. Written consent from all participants was obtained, as stated in the ethics statement. Participants were provided with a consent form that clearly explained the study’s purpose, procedures, potential risks, and their rights as participants. The consent form was written in a language that participants understood and they were given sufficient time to review and ask any questions before providing their written consent. In cases where verbal consent was obtained from illiterate respondents, we have documented and witnessed it according to the established protocol.

A total of 50 subjects were recruited in each EB. All recruited subjects had a brief interview, where demographic, medical and ocular history data were taken. It was followed by visual acuity assessment at a distance of three meters using tablets installed with the RAAB7 application with built-in quality control measures [7,8]. An eye examination was performed by the doctors in the survey team using a hand-held ophthalmoscope. Should subjects have a visual impairment, the primary cause was identified, and the subjects were referred to the nearest ophthalmic care facility for further management.

Inter-observer variation was conducted separately in each region before the survey. During the inter-observer variation assessment, six survey teams were assessed in their agreements, asking 15 questions to subjects comprising patients and staff from the Ophthalmology Outpatient Department. There were 20 subjects with mixed normal and impaired vision, including cataracts, (pseudo)aphakia or posterior segment diseases. The six survey teams examined the same 20 individuals during the assessment. The other teams’ findings were compared to those of the most senior or experienced team (as the gold standard).

### Definition

National Eye Survey, NES II, was conducted in 2014, and the survey under discussion, NES III, was conducted in 2023. Although the later survey only involved two out of six regions, the national PBL committee agreed to maintain the “National/NES” label for branding and advocacy purposes.

The Kappa coefficient values were classified as poor (*<*0.20), fair (0.21-0.40), moderate (0.41-0.60), good (0.61-0.80), and very good (0.81-1.00).

The prevalence of distance VI was reported using Presenting Visual Acuity (PVA) in the better eye, measured with correction, if available. VI categories were defined according to the Visual Acuity (VA) thresholds used in the WHO’s International Classification of Diseases (ICD-11) [9]:

- Blindness: PVA less than 3/60 in the better eye
- Severe VI: PVA less than 6/60 to 3/60 in the better eye
- Moderate VI: PVA less than 6/18 to 6/60 in the better eye
- Mild VI: PVA less than 6/12 to 6/18 in the better eye

Once identified after eye examination, the primary cause of VI was categorized into treatable, preventable, avoidable, and posterior segment disease. Treatable VI were conditions that can be treated (for example, Refractive Error, Aphakia, and Cataract). Preventable VI were conditions that could be prevented by Primary Health Care (PHC), Primary Eye Care (PEC), or specialized ophthalmic services. Treatable and Preventable constituted Avoidable.

### Sample size calculation

The latest population data was obtained from the Malaysian National Census 2020 [10, 11]. A prevalence of blindness of 1.5 % in Eastern and 1.6% in Sarawak among subjects aged 50 and older from NES II (2014) was used in the calculation using a 95% confidence interval, precision of 30% and estimated design effect (DEFF) of 1.5. It took into consideration the possibility of 20% non-response [4,5]. The calculation resulted in a sample size of 105 clusters (5239 subjects aged 50 years and older) and 98 clusters (4900 subjects aged 50 years and older) for Eastern and Sarawak, respectively.

### Statistical Analysis

Data were entered into the cloud-based RAAB7 software using tablets. The software had an in-built inconsistency check and validation for double data entry. It reported the prevalence of VI in percentages and 95% Confidence Interval values by adjusting for age and sex. Other categorical data were reported in frequency and percentage. Raw data and digital reports were generated automatically in real-time and accessible to the authorized investigators through the web-based portal.

## Results

The Kappa results for the Eastern survey team were 24.0% moderate, 50.7% good and 25.3% very good agreement with the “gold standard” survey team. The Sarawak survey team were 1.3% fair, 34.7% moderate, 40.0% good and 24.0% very good agreement with the “gold standard” survey team.

A total of 10,184 subjects 50 years old and older were enumerated: Eastern, n= 5,250, Sarawak, n= 4,934). A total of 9,709 were examined: Eastern, n= 4,961 (94.5% response), Sarawak, n= 4,748 (96.2% response). These subjects represented 0.5% of all people aged 50 and older in both regions. Of the 475 non-respondents, 73 (0.7%) refused to participate, 329 (3.3%) were incapable of being examined due to communication problems such as deafness or dementia, and 73 (0.8)% were not available. More female subjects were examined, n= 5,520 (56.8%) (Table 1).

**Table 1.** Examination Status, NES III (2023).

| **EASTERN** | Female |  | Male |  | Total |  |
| --- | --- | --- | --- | --- | --- | --- |
| **Examination Status:** | n | % | n | % | n | % |
| Examined* | 2829 | 95.6 | 2132 | 93 | 4961 | 94.5 |
| Refused | 20 | 0.7 | 17 | 0.7 | 37 | 0.7 |
| Incapable | 83 | 2.8 | 105 | 4.6 | 188 | 3.6 |
| Unavailable | 26 | 0.9 | 38 | 1.7 | 64 | 1.2 |
| Total** | 2958 | 100 | 2292 | 100 | 5250 | 100 |
| **SARAWAK** | Female |  | Male |  | Total |  |
| **Examination Status:** | n | % | n | % | n | % |
| Examined* | 2691 | 96.4 | 2057 | 96 | 4748 | 96.2 |
| Refused | 28 | 1 | 8 | 0.4 | 36 | 0.7 |
| Incapable | 67 | 2.4 | 74 | 3.5 | 141 | 2.9 |
| Unavailable | 6 | 0.2 | 3 | 0.1 | 9 | 0.2 |
| Total** | 2792 | 100 | 2142 | 100 | 4934 | 100 |

\* Total examined (Percentage is the response rate)
\*\* Total enumerated

The sample had less younger subjects in both genders (50-59 years old) than the survey area, but had more subjects in the older age group (except in the age group 80+ in Sarawak) (Fig 2). All the results, therefore, were analyzed using age-sex-adjusted estimates due to the difference.

**Fig 2.**
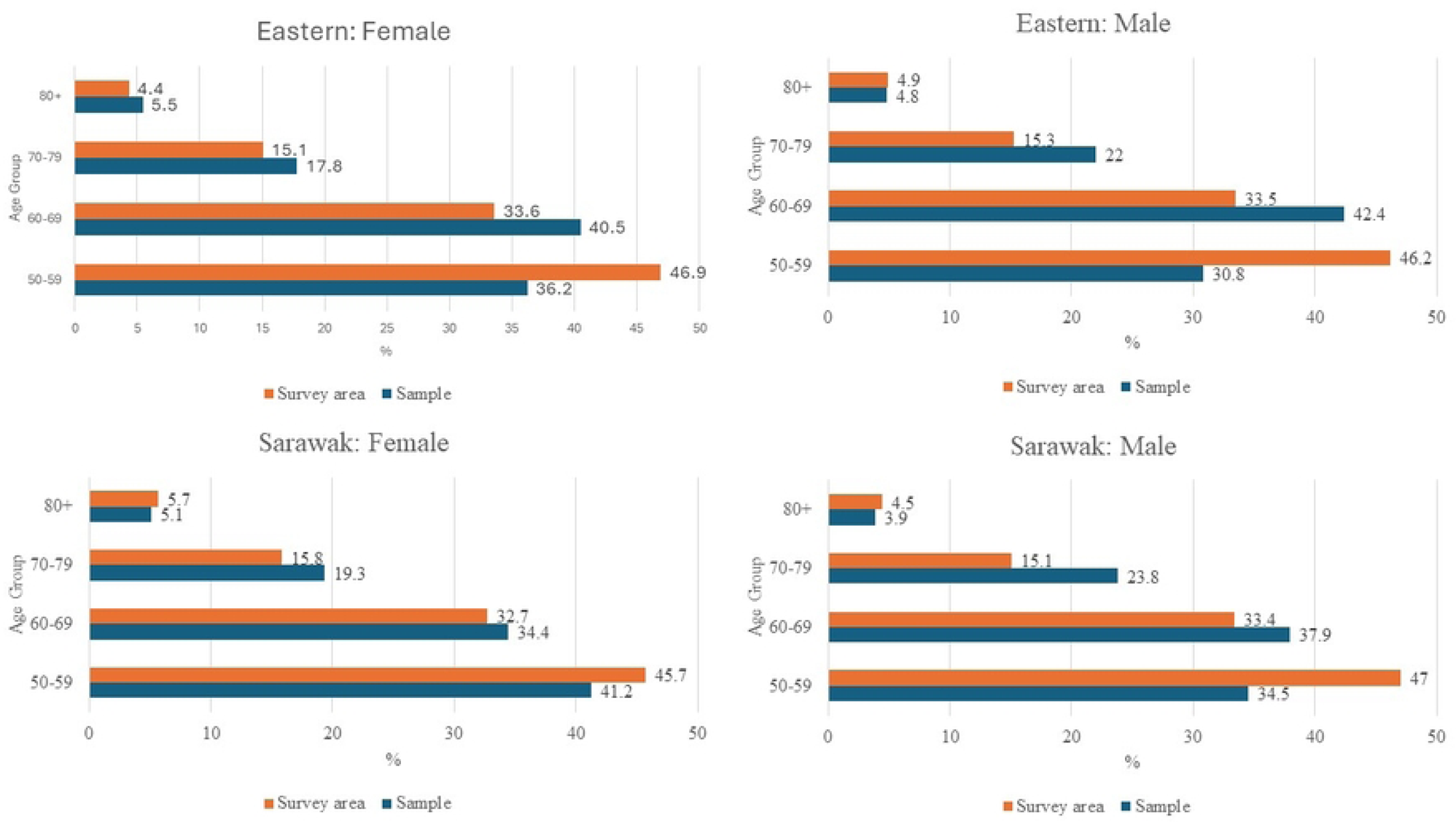
Population in Sample vs Survey Areas, NES III (2023).

There was a higher prevalence of blindness in the Eastern Region than Sarawak: 0.8%, 95%CI (0.5, 1.1) vs 0.6%, 95%CI (0.3,0.9). However, the prevalence of blindness and prevalence of severe VI appeared to decrease, comparing the results to NES II (2014): Blindness Eastern from 1.4%, 95%CI (0.9, 1.9) to 0.8%, 95%CI (0.5, 1.1) and Blindness Sarawak from 1.6% 95%CI (1.0, 2.1) to 0.6%, 95%CI (0.3, 0.9). Severe VI Eastern from 1.2%, 95%CI (0.8, 1.7) to 0.9%, 95%CI (0.6, 1.1) and Severe VI Sarawak from 1.1% 95%CI (0.6, 1.6) to 0.9%, 95%CI (0.6, 1.2) (Table 2). There was no significant difference between males and females in any category of Visual Impairment during both surveys (Table 2).

**Table 2.**
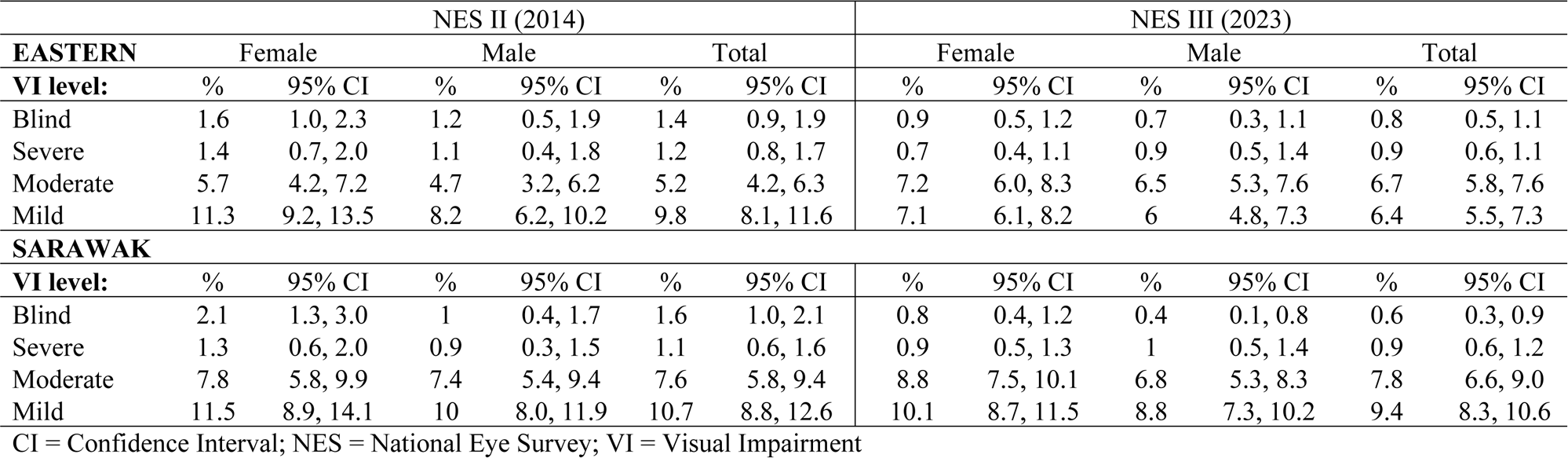
Adjusted prevalence and magnitude of Blindness, severe, moderate, and mild vision impairment-Comparing NES II (2014) and NES III (2023).

The main cause of blindness was untreated cataracts: 77.3% (Eastern) and 75.0% (Sarawak). Although diabetic retinopathy was the 2^nd^ leading cause of blindness in Eastern (9.1%) and severe VI (6.2%), it was not the cause of blindness but the 4^th^ leading cause of severe VI in Sarawak (2.1%) (Table 3). As high as 95.5% of the primary causes of blindness in Eastern and 90.6% in Sarawak were avoidable (Table 4).

**Table 3.** Principal cause of blindness, severe, moderate, and mild vision impairment, NES III (2023).

| EASTERN |  |  |  |  |  |  |  |  |  | SARAWAK |  |  |  |  |  |  |  |
| --- | --- | --- | --- | --- | --- | --- | --- | --- | --- | --- | --- | --- | --- | --- | --- | --- | --- |
|  |  | Blind |  | Severe |  | Moderate |  | Early |  | Blind |  | Severe |  | Moderate |  | Early |  |
| No | Principal.Cause | n | % | n | % | n | % | n | % | n | % | n | % | n | % | n | % |
| 1 | Refractive error | 0 | 0 | 2 | 4.2 | 182 | 46.4 | 287 | 79.3 | 0 | 0 | 2 | 4.3 | 148 | 35.7 | 307 | 64 |
| 2 | Aphakia | 0 | 0 | 0 | 0 | 0 | 0 | 0 | 0 | 0 | 0 | 0 | 0 | 0 | 0 | 0 | 0 |
| 3 | Cataract | 34 | 77.3 | 39 | 81.2 | 179 | 45.7 | 63 | 17.4 | 24 | 75.0 | 38 | 80.9 | 246 | 59.3 | 143 | 29.8 |
| 4 | Cataract surgical complications | 0 | 0 | 0 | 0 | 4 | 1 | 0 | 0 | 0 | 0 | 1 | 2.1 | 2 | 0.5 | 3 | 0.6 |
| 5 | Trachomatous corneal opacity | 0 | 0 | 0 | 0 | 0 | 0 | 0 | 0 | 0 | 0 | 0 | 0 | 0 | 0 | 0 | 0 |
| 6 | Other corneal opacity | 0 | 0 | 1 | 2.1 | 1 | 0.3 | 0 | 0 | 1 | 3.1 | 0 | 0 | 0 | 0 | 0 | 0 |
| 7 | Pterygium | 1 | 2.3 | 0 | 0 | 5 | 1.3 | 3 | 0.8 | 1 | 3.1 | 2 | 4.3 | 8 | 1.9 | 10 | 2.1 |
| 8 | Phthisis | 1 | 2.3 | 0 | 0 | 0 | 0 | 0 | 0 | 2 | 6.2 | 0 | 0 | 0 | 0 | 0 | 0 |
| 9 | Onchocerciasis | 0 | 0 | 0 | 0 | 0 | 0 | 0 | 0 | 0 | 0 | 0 | 0 | 0 | 0 | 0 | 0 |
| 10 | Glaucoma | 2 | 4.5 | 1 | 2.1 | 7 | 1.8 | 0 | 0 | 1 | 3.1 | 2 | 4.3 | 1 | 0.2 | 3 | 0.6 |
| 11 | Diabetic Retinopathy | 4 | 9.1 | 3 | 6.2 | 8 | 2 | 6 | 1.7 | 0 | 0 | 1 | 2.1 | 7 | 1.7 | 10 | 2.1 |
| 12 | Age-related macular degeneration | 0 | 0 | 0 | 0 | 3 | 0.8 | 1 | 0.3 | 0 | 0 | 0 | 0 | 1 | 0.2 | 0 | 0 |
| 13 | Other posterior segment disease | 2 | 4.5 | 2 | 4.2 | 3 | 0.8 | 2 | 0.6 | 3 | 9.4 | 1 | 2.1 | 2 | 0.5 | 3 | 0.6 |
| 14 | Myopic degeneration | 0 | 0 | 0 | 0 | 0 | 0 | 0 | 0 | 0 | 0 | 0 | 0 | 0 | 0 | 0 | 0 |
| 15 | Other globe or CNS abnormalities | 0 | 0 | 0 | 0 | 0 | 0 | 0 | 0 | 0 | 0 | 0 | 0 | 0 | 0 | 1 | 0.2 |
| Total |  | 44 | 100 | 48 | 100 | 392 | 100 | 362 | 100 | 32 | 100 | 47 | 100 | 415 | 100 | 480 | 100 |

CNS = Central Nervous System

**Table 4.** Principal cause of Blindness (PVA <3/60), by gender and intervention category, NES III (2023).

| EASTERN |  | Male |  | Female |  | Total |  |
| --- | --- | --- | --- | --- | --- | --- | --- |
| Group | Intervention Category (refer Table 3 for numbers) | n | % | n | % | n | % |
| A | Treatable (1, 2, 3) | 22 | 81.5 | 12 | 70.6 | 34 | 77.3 |
| B | Preventable (PHC/PEC services) (5, 6, 7, 8, 9) | 1 | 3.7 | 1 | 5.9 | 2 | 4.5 |
| C | Preventable (Ophthalmic services) (4, 10, 11) | 3 | 11.1 | 3 | 17.6 | 6 | 13.6 |
| D | Avoidable (A + B + C) | 26 | 96.3 | 16 | 94.1 | 42 | 95.5 |
| E | Posterior segment disease (9, 10, 11, 12, 13, 14) | 4 | 14.8 | 4 | 23.5 | 8 | 18.2 |
| SARAWAK |  |  |  |  |  |  |  |
| Group | Intervention Category (refer Table 3 for numbers) | n | % | n | % | n | % |
| A | Treatable (1, 2, 3) | 17 | 77.3 | 7 | 70.0 | 24 | 75.0 |
| B | Preventable (PHC/PEC services) (5, 6, 7, 8, 9) | 3 | 13.6 | 1 | 10.0 | 4 | 12.5 |
| C | Preventable (Ophthalmic services) (4, 10, 11) | 1 | 4.5 | 0 | 0 | 1 | 3.1 |
| D | Avoidable (A + B + C) | 21 | 95.5 | 8 | 80.0 | 29 | 90.6 |
| E | Posterior segment disease (9, 10, 11, 12, 13, 14) | 2 | 9.1 | 2 | 20.0 | 4 | 12.5 |

PVA = Presenting Visual Acuity; PHC = Primary Health Care; PEC = Primary Eye Care

## Discussion

The availability of RAAB7 software facilitated data entry and cleaning during NES III. The web platform with live survey data visibility enabled real-time monitoring for quality assurance throughout the survey. The trainer and investigators could identify and respond to any issues and questions from the data collectors on the ground in real time. Compared to NES II (2014), digitalization of data entry, cleaning, analysis and delivery of reports during NES III (2023) assured data quality. It saved time and, therefore, costs needed to support the survey teams on the ground as the duration of data collection was shortened.

Females formed 56.4% out of the 10,184 subjects enumerated. This population gender ratio was not consistent with the national census [10,11]. The sample also had less younger subjects in both genders (50-59 years old) than the survey area, but had more subjects in the older age group (except in the age group 80+ in Sarawak). It could be explained by the possibility that females in both regions tend to stay at home, not working, resulting in them having a higher chance of being enumerated. The younger age group (50-59 years old) in both genders could also likely be away at work when the survey teams visited due to work or social commitments. This pattern is consistent and can be seen in other surveys [12–16].

The prevalence of blindness during the six simultaneous RAAB (NES II 2014) in Malaysia ranged from 0.5-1.9% (with a calculated weightage of 1.2%) [4]. The prevalence was reduced to 0.8% and 0.6% for two regions (Eastern and Sarawak, respectively) during NES III in 2023. In general, the prevalence of VI during NES II and III were lower compared to other countries in the South East Asia Region [17–19]. It is essential to acknowledge the country’s position regarding the prevalence compared to neighbouring countries, but achieving a reduction following an intervention is more important or meaningful.

The prevalence of diabetic retinopathy resulting in visual impairment in Eastern was higher than in Sarawak. This is consistent with the findings of a higher prevalence of diabetes mellitus in Eastern than Sarawak [20]. We acknowledge that more work needs to be done at the Ministerial level with the Non-Communicable Disease division to strategically collaborate to curtail the increasing number of people with diabetes mellitus and prevent diabetic retinopathy through more active nationwide screening initiatives.

One of the national initiatives which can potentially be strengthened for diabetic retinopathy screening is the *Klinik Katarak-Kementerian Kesihatan Malaysia* (Cataract Clinic Ministry of Health Malaysia, *KK-KKM*), an outreach arm of the ministry to reach the population (Fig 3). It was launched in 2014 in Sarawak and the Eastern Region of Malaysia as part of the country’s progress commitments with the World Health Assembly (WHA) 66.4 resolution. It was also implemented as part of the national action plan following the findings of the NES II (2014). The modified buses transport surgical and medical equipment along the selected service routes according to the location of Provincial Hospitals, which are used as the primary service sites. Once arrived at the site, the equipment is offloaded and used in the clinic (for screening) or operating room for (cataract surgery).

**Fig 3.**
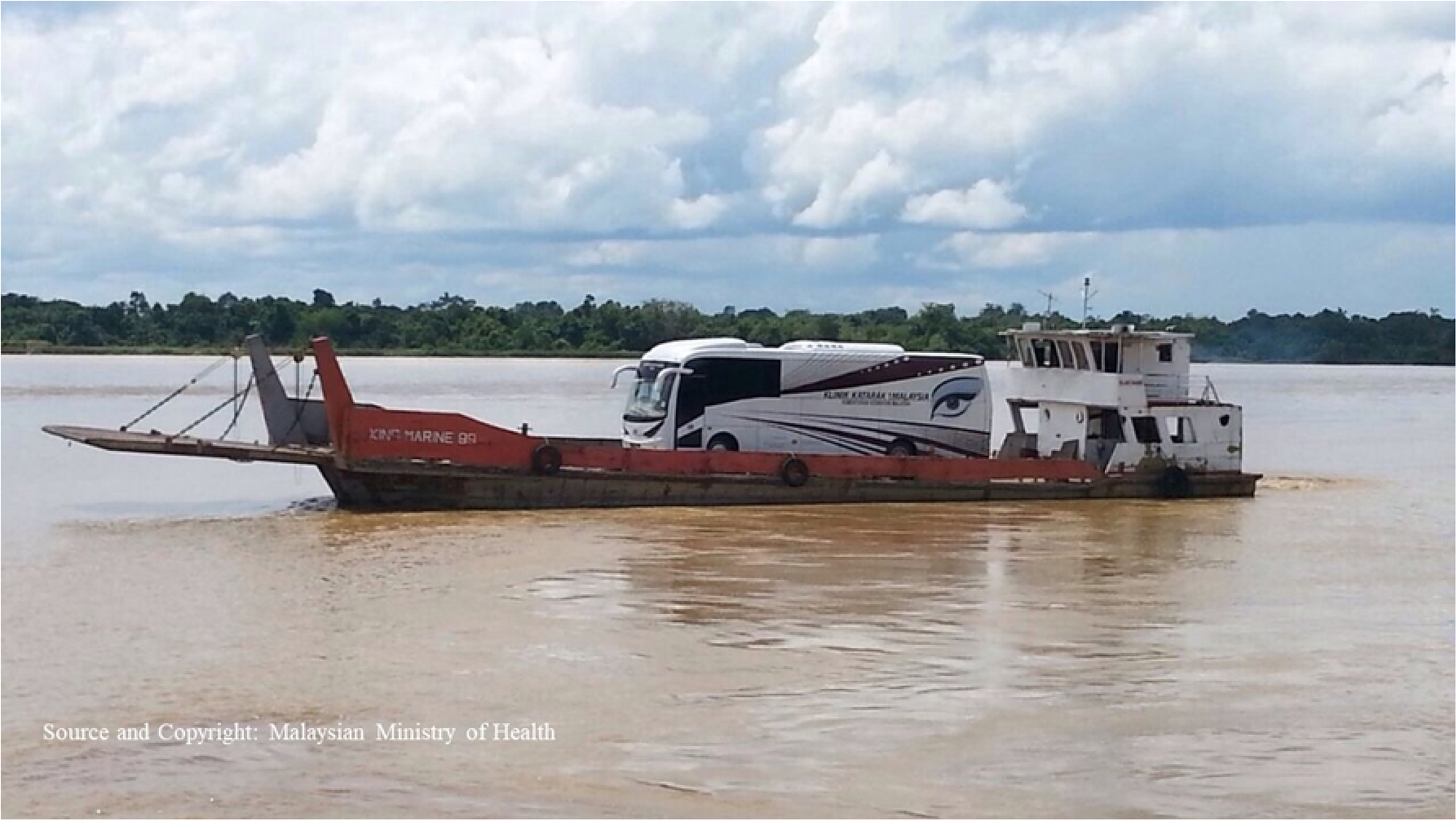
The Mobile Unit on a boat en route to Kapit, a remote district in Sarawak.

The service concept is based on operating near patients’ homes. Unlike other cataract mobile units in other countries, surgery is not done on the bus [21–23]. Instead, all activities optimize the provincial hospitals’ existing facilities that otherwise have low usage due to low population density. The KK-KKM project in both regions is focused on scheduled trips for screening, surgery and revisiting after one month to assess visual outcomes. The timetable is distributed to all the provincial hospitals at the beginning of the calendar year. The fixed schedule allows people in remote areas to plan their finances and trips to come forward and seek eye treatment.

Although branded as a cataract surgical program, the screening platform and community engagement activities within the program encourage eye care advocacy within the community. KK-KKM allows screening of any individuals with any eye problems. They will be treated or referred to the nearest hospitals accordingly. It features a collaboration of multiple stakeholders and eye care providers from the community, Non-Governmental Bodies (NGOs), local Councils, Community Nurses, Primary Eye Care Doctors, Optometrists and Ophthalmologists. Data from cataract surgeries performed at the KK-KKM locations are also monitored centrally for the centre and individual surgeons’s performance monitoring [24–31].

The concept of “Bringing High Impact Quality Eye Care Closer to Home“, community engagement/advocacy and performance monitoring in outreach cataract surgery could have explained the reduction in the prevalence of blindness within both regions after nine years of service. The objective, concept and work process were endorsed by WHO when it was selected as a Case Study for the Western Pacific WHO Innovation Challenge in 2021/2022 [32].

## Conclusion

The prevalence of blindness and severe VI were reduced. The reduction could have been attributed to the KK-KKM, an outreach project operational at the community level. However, more efforts are needed to address the high percentage of avoidable blindness within both regions.

## Data Availability

All survey data are available from the database URL https://www.raab.world/survey-data

## Acknowledgments

The authors would like to thank the Director General of the Ministry of Health Malaysia for his permission to publish this article. The authors would also like to acknowledge the contribution of data entry by the data collectors during both NES II and III.

## Notes

### Competing Interest Statement

The authors have declared no competing interest.

### Funding Statement

Yes

### Author Declarations

Medical Research and Ethics Committee of the Malaysian Ministry of Health (Research ID NMRR-19-197-46172). The study was conducted in accordance with the tenets of the Declaration of Helsinki.

